# Association between older patients receiving geriatric co-management at the emergency department and acute hospital admissions compared to usual care: an observational, controlled study

**DOI:** 10.1101/2025.09.03.25334977

**Authors:** Vera M. Hogervorst, Janet L. MacNeil Vroomen, Marthe Ribbink, Bianca M. Buurman, Annemarieke De Jonghe

**Affiliations:** Department of Internal Medicine, Section of Geriatric Medicine, Amsterdam Public Health Institute, Amsterdam UMC, University of Amsterdam, Amsterdam, The Netherlands; Department of Geriatric Medicine, Tergooi MC, Hilversum, The Netherlands

**Keywords:** Older people with frailty, Emergency department, Comprehensive geriatric assessment, Observational study, Hospital admissions, Geriatric co-management, Geriatric emergency medicine, GEM-team

## Abstract

**Objectives:** The aim of this study is to determine if a geriatric co-management model, referred to as “The Geriatric Emergency Medicine (GEM)-team’ is associated with less admissions to hospital in older patients compared to the usual care without increasing the risk of mortality or 30-day ED readmissions.

**Design:** This observational, controlled study used 18-month data prospectively collected from hospital records. Inverse probability weighted, logistic and linear regression models were used for evaluation.

**Setting:** An Emergency Department at a suburban Dutch, general hospital, receiving approximately 10,000 patients aged 70 or older.

**Participants:** All patients aged 70 or older were screened according to predefined criteria. When positively screened patients were presented at the ED on weekdays between 9AM and 5PM they received geriatric co-management. Outside these hours and when capacity of the GEM-team was reached, patients received care as usual.

**Interventions:** Geriatric co-management at the ED, involves a geriatric multidisciplinary team in collaboration with the primary ED-physician who share management and responsibility for the provided medical treatment and nursing care starting directly at the primary assessment.

**Primary and secondary outcome measures:** The primary outcome was hospital admission and secondary outcomes were the composite outcome of 30-day ED readmissions and mortality.

**Results:** Patients receiving geriatric co-management (n=972) had lower odds for hospitalization after adjusting for the propensity score (odds ratio [OR]: 0.61, 95% confidence interval [CI]: 0.50-0.75) compared to the control-group (n=1355) while 30-day ED readmissions and mortality did not differ between groups (OR 0.95, 95% CI: 0.75-1.22).

**Conclusions:** Geriatric co-management at the ED is associated with decreased hospital admissions while 30-day ED readmissions or mortality was not impacted. These preliminary results contribute to the evidence that geriatric co-management may be an effective intervention for older patients with frailty at the ED.

## INTRODUCTION

Emergency departments (ED) are challenged by the multiple needs of the growing population of older patients with frailty(1), as the ED is originally designed to take care of single acute conditions such as heart attacks, fractures or infections(2, 3). Older patients are frequently acutely admitted to hospital, which is associated with negative outcomes like mortality, long length of stay and discharges to other destinations than home (4, 5). Taking care of older patients living with frailty is complex because of non-specific and atypical presentation of illness, multiple co-morbidities, polypharmacy, functional decline and altered homeostasis (2, 6). Diagnostic challenges arise because of communication difficulties such as impaired memory functions, loss of hearing or speech and the need to communicate with informal caregivers (7). The complexity of care increases because of existential considerations related to altered perspectives on sickness and health in the light of the end-of-life nearing (8, 9). Even older patients who have acquired a minor injury, have a risk for an unneeded, undesired, and costly hospital admission because of all other patient-related factors (10, 11).

Interventions targeted at older patients at the ED have been introduced in different health care systems over the world with variations within the exact models and resourcing (12–15). A holistic, multi-domain assessment used by geriatric trained healthcare professionals called Comprehensive Geriatric Assessment (CGA) is quite often included(16–18), sometimes combined with a discharge program or a medication management program. Evidence suggests that multi-strategy interventions, with all three components present, may be associated with reducing hospital admissions (19). Providing CGA at the ED is difficult because of the ED time-pressures, costs and the increasing numbers of patients living with frailty (20). Some intervention models have initiated geriatric EDs where a CGA is provided by a dedicated team (21). Other interventions provide a CGA by consulting geriatric trained nurses who perform secondary geriatric screening by using pre-specified screening lists (22). Many hospitals face budget restrictions and therefore do not have access to provide costly services as geriatric EDs(23). The evidence on how to provide a geriatric assessment at the ED is scarce(24), however, a CGA applied to the urgent care setting, is generally accepted as the best way to assess older patients living with frailty at the ED (18).

Geriatric co-management uses the joint expertise of geriatric trained healthcare professionals and the primary physician and is implemented in clinical settings like orthopedics, oncology and cardiology (25). They share responsibility for treatment plans, discharge coordination and clinical outcomes. Geriatric co-management brings added resources and assistance, geriatric expertise and working relations with regional care partners (26). It is associated with reduced length of stay and complications throughout hospital admission (25, 27). In The Netherlands geriatric co-management applied at the ED is novel and has not been evaluated.

A geriatric co-management model called the Geriatric Emergency Medicine (GEM)-team was initiated at a general Dutch hospital. The team is staffed by either an Advanced Nurse Practitioner (ANP) or medical doctor (MD) trained in Geriatric Emergency Medicine. Within the time window the ED-physician needs to assess the patient, the GEM-team simultaneously provides their integrated medical and nursing CGA work-up. Special attention is given to non-specific and atypical presentation of illness, co-morbidities, polypharmacy with the help of a pharmacy team, advanced care directives and shared decision making. The ED-physician, GEM-team ANP/MD and the patient decide together if and what kind of treatment is needed and where the treatment and care should take place. The GEM-team also involves liaison nurses who organize direct transfer of the patient from the ED to home or an intermediate care facility when indicated.

The aim of this study is to evaluate if a geriatric co-management model, referred to as ‘The GEM-team’ is associated with less acute admissions to hospital in older patients compared to usual care without increasing the risk of mortality or 30-day ED readmissions.

## METHODS

### Study design

This observational, controlled study evaluated patients that were presented to the ED and co-managed by the GEM-team compared to a control group who received usual care for an eighteen-month period. Anonymized data retrieved from the electronic hospital records (EHR) was used for this study. The STROBE (28) and TIDieR guidelines (additional file 1) were used for reporting.

### Setting

In May 2020, the GEM-team was implemented between 9AM and 5PM on weekdays, on a twenty bed ED at one of the two locations of a general teaching hospital located in a suburban area in The Netherlands. This ED served patients with surgical, orthopedic, or internal medicine problems. Within the catchment area of the hospital no other hospital exists. During the study period no short stay area was available for ED patients. In 2018 a regional covenant between the hospital and regional intermediate care facilities was established. The covenant describes the agreements and logistics regarding transfer of ED-patients to an intermediate care facility at all hours.

### Participants and intervention

All patients aged 70 or older who visited the ED during the eighteen-months’ time-window were triaged at presentation by the ED-nurse. The ED-nurse then applied the GEM-score to decide if geriatric co-management was indicated. The GEM-score quickly screens for the presence of cognitive problems, delirium, fall/collapse-related presentation, or anticipated discharge problems. The last item of the GEM-score was the opinion of the ED-nurse whether the patient would benefit from geriatric co-management. Cognitive problems were screened for by reviewing medical notes or hand-over for a recorded cognitive disorder diagnosis or by verbal confirmation of next-of-kin. The presence of delirium was assessed by an ED-nurse and if needed, a confirmation of the next-of-kin about a recent change in behavior of the patient. The GEM-score has a binary outcome, yes or no. If one or more items of the GEM-score was positively confirmed, the GEM-team started their geriatric co-management trajectory. While the GEM-score is not a validated tool, it is based on validated scores as the VMS-score, APOP-score and ISAR HP(29) and a pragmatic expert opinion of the Geriatric and Emergency Department team at the hospital. The inclusion of positively GEM-score screened patients in the GEM-cohort was mainly time-dependent, but also influenced by the availability of the Advanced Nurse Practitioner (ANP) or medical doctor (MD) of the GEM-team. When too many GEM-patients were presented at once at the ED, inclusion in the GEM-cohort was left to the discretion of the GEM-ANP/MD. Patients with high risk for unnecessary hospital admission and/or considered highly frail determined by the Clinical Frailty Score (30) were then favored.

The GEM-team is a collaboration of an ANP or MD trained in GEM; ED-nurses; a geriatrician; a pharmacy team and liaison nurses. In the Netherlands, ANPs are senior nurses with a master’s degree and 2 years of medical training into a pre-specified medical expertise area. They are legally authorized to autonomously provide integrated medical and nursing assessments (31). The MD is supervised by the geriatrician and was provided with on-site training regarding assessing older patients at the ED at our teaching hospital. The ANP liaises with the geriatrician when the assessment of the patient exceeds their medical expertise. Both the ANP and the MD provide an integrated medical and nursing geriatric assessment based on the CGA-framework adapted to the urgent care setting (18) in co-management with the primary ED-physician. The GEM-team is based at the ED and was assigned to perform CGA within the time-window the primary ED-physician needed to assess the patient. The GEM-team complements the primary ED-physician starting at the primary assessment. They decided together which laboratory tests, imaging and consults were needed. The GEM-team ANP/MD considered treatment preferences and advance care directive of the patient. Special attention was given to non-specific complaints and atypical presentation of illness, co-morbidities, functional decline and altered homeostasis, as these are common pitfalls known in the assessment of older patients. Polypharmacy and drug-related ED-presentations were assessed together with the pharmacy team(32). The GEM-team ANP/MD collaborated with the ED-nurses regarding secondary geriatric nursing screens, investigating the social network of the patient, indicating preventive measures, and providing a geriatric friendly stay at the ED. When all tests and consultations have taken place, a quick mini multidisciplinary discussion is being held which further enrols into talking with the patient and next of kin. Treatment options, advanced care directives and the location of the treatment and care (hospital, permanent/intermediate care facility or at home) are considered. When treatment and care at a permanent or intermediate care facility was chosen or district nursing at home, the hospital-based liaison nurses organized the transfer in collaboration with the regional healthcare providers. When hospital treatment is not indicated or desired, preference was to transfer patient home or to a permanent or intermediate care facility. At discharge the GEM-team ANP/MD provided the patient and next of kin with discharge information and follow-up appointments. A written hand-over and/or telephone call with the regional healthcare providers is conducted at discharge of the ED. At admission to the hospital, treatment instructions were written at the Electronic Health Records (EHR) and verbally handed over. If indicated a geriatric co-management trajectory throughout hospital admission was initiated (Figure 1).

**Figure 1.**
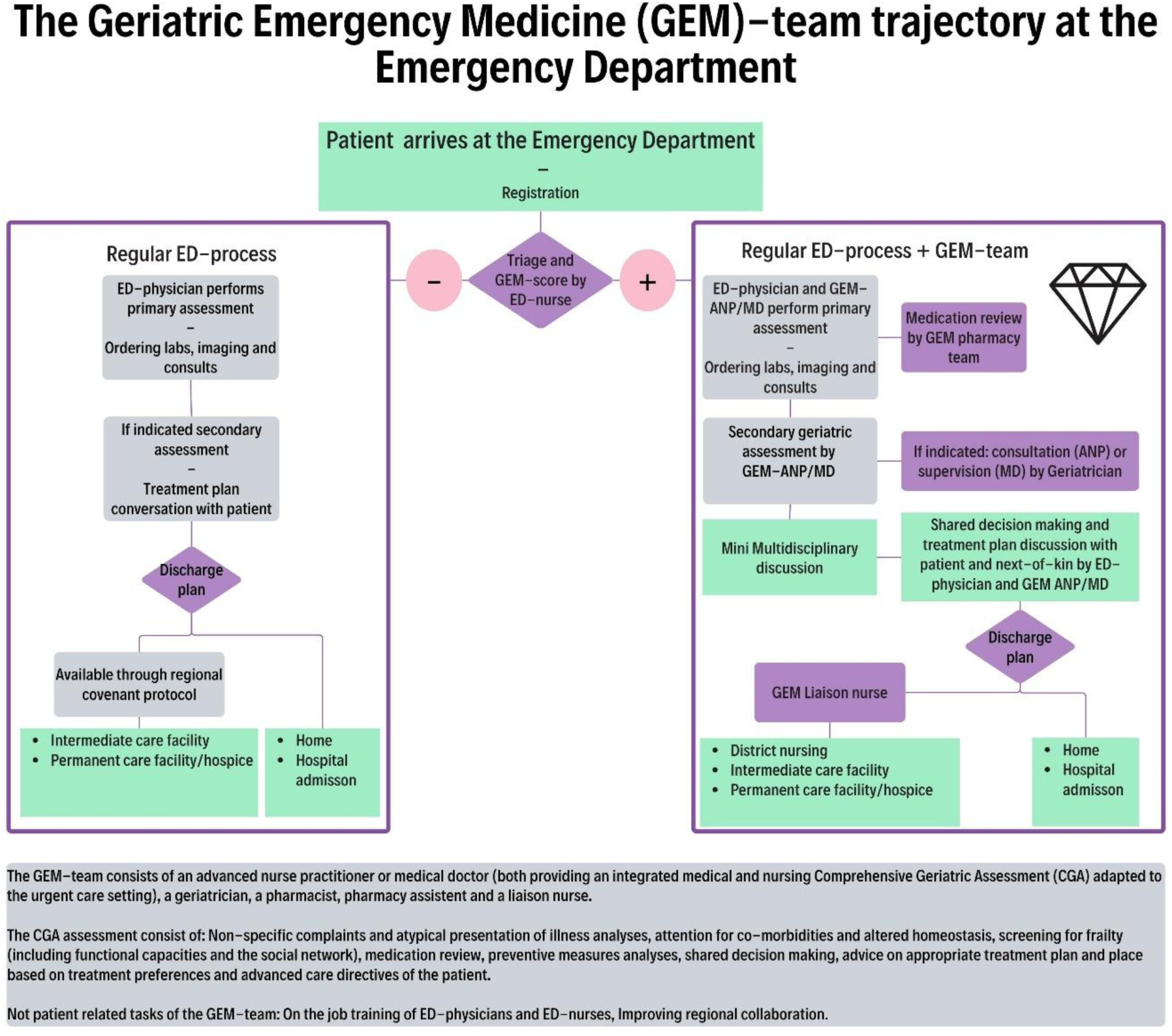
The Geriatric Emergency Medicine (GEM)-team trajectory at the Emergency Department.

For this study two cohorts were composed of all consecutively included patients aged 70 or older presented at the ED between May 2020 and November 2021. All patients not screened with the GEM-score or screened with the GEM-score but scored negatively were excluded. Patients screened positively at the GEM-score and co-managed by the GEM-team formed the GEM-cohort and patients screened positively but not seen by the GEM-team formed the control-cohort (Figure 2) This cohort received care as usual including the possibilities created with the regional covenant.

**Figure 2.**
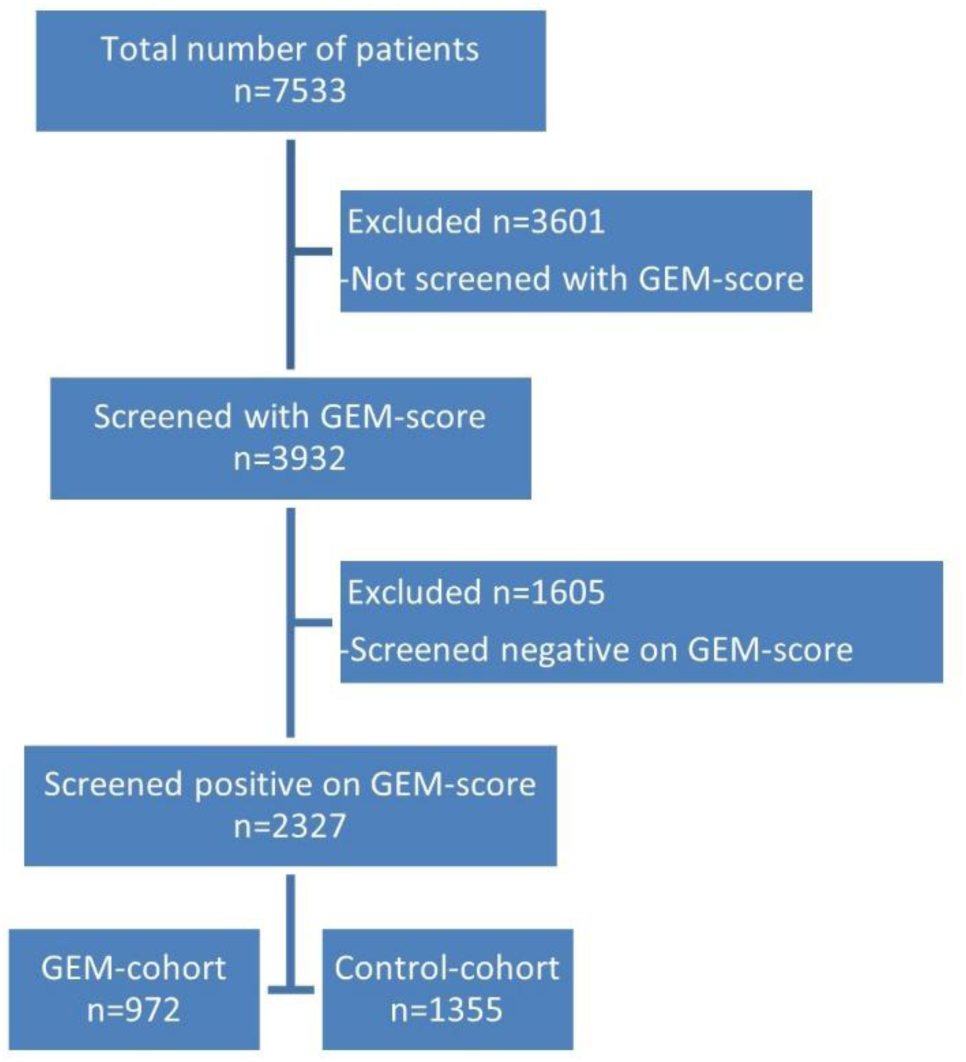
Enrolment of patients, 70 years and older, presented at the Emergency Department.

### Data Collection

Eighteen-months of data were prospectively collected at the EHR of the patient. As part of the CGA the Clinical Frailty Scale was assessed (33) and discharge settings and/or destinations were noted. Data was anonymously retrieved from the EHR and municipal records with the help of administrative staff. Baseline data included gender, age at presentation, date and time of ED presentation, triage urgency, medical specialty of the primary ED-physician. Collected outcome data included date of death, admission to hospital and discharge dates and 30-day ED-readmission. All data was uploaded in SPSS 28 and R version 4.0 for further analyses. Missing data regarding baseline characteristics was recorded in table 1 and accepted. Outcome data was available and calculated for all patients. Data checks were performed. Acute admission to hospital was defined as: being transferred from the ED to a clinical ward or theatre at the hospital and having a hospital stay larger than 24 hours (including the hours at the ED). ED-admissions have never exceeded 24 hours. All members of the GEM-team and some investigators based at the general hospital had access to parts of the EHR. To minimize chances of bias, it was chosen to perform the analyses on the anonymized data files in collaboration with external investigators based at the academic hospital.

**Table 1.**
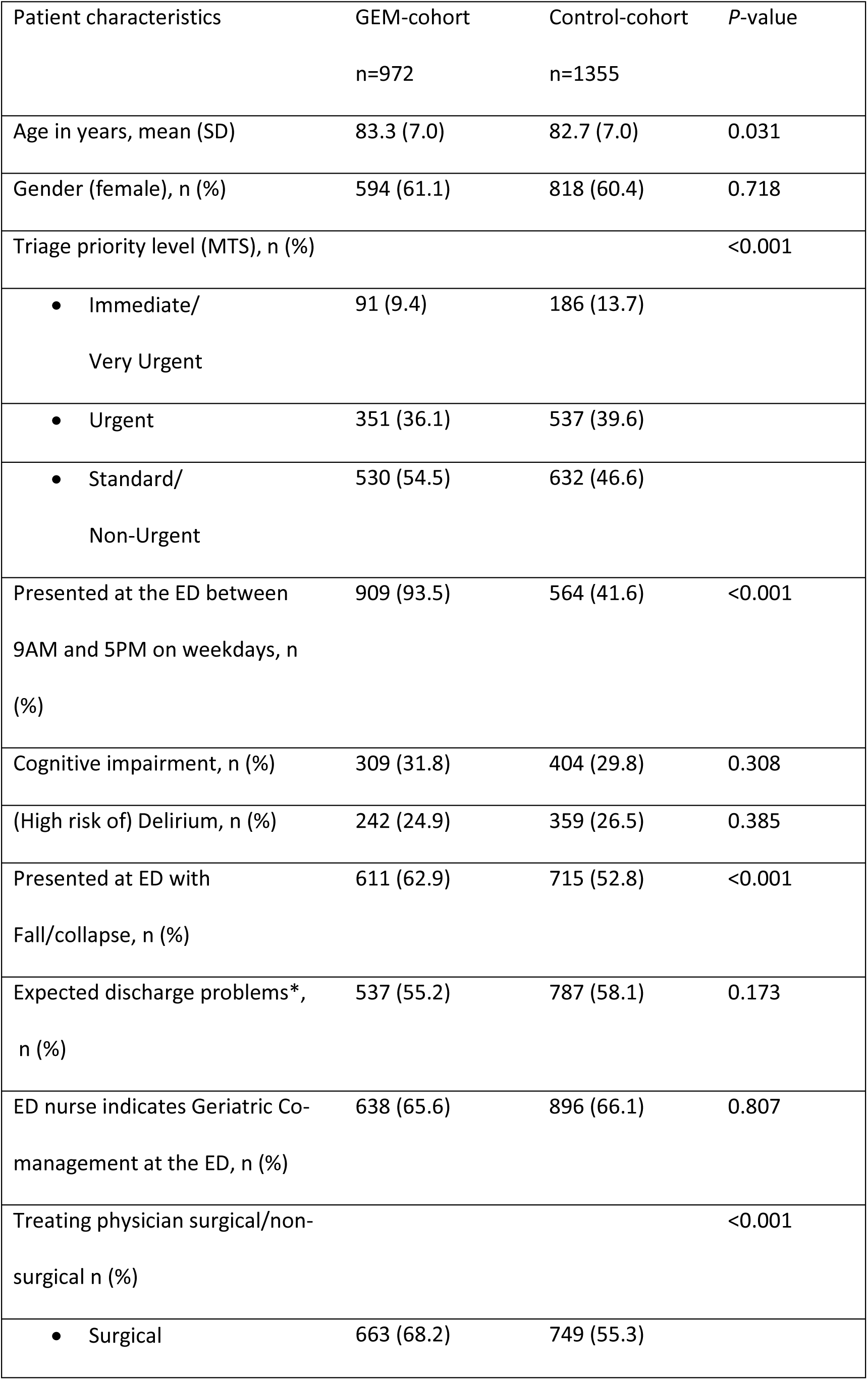
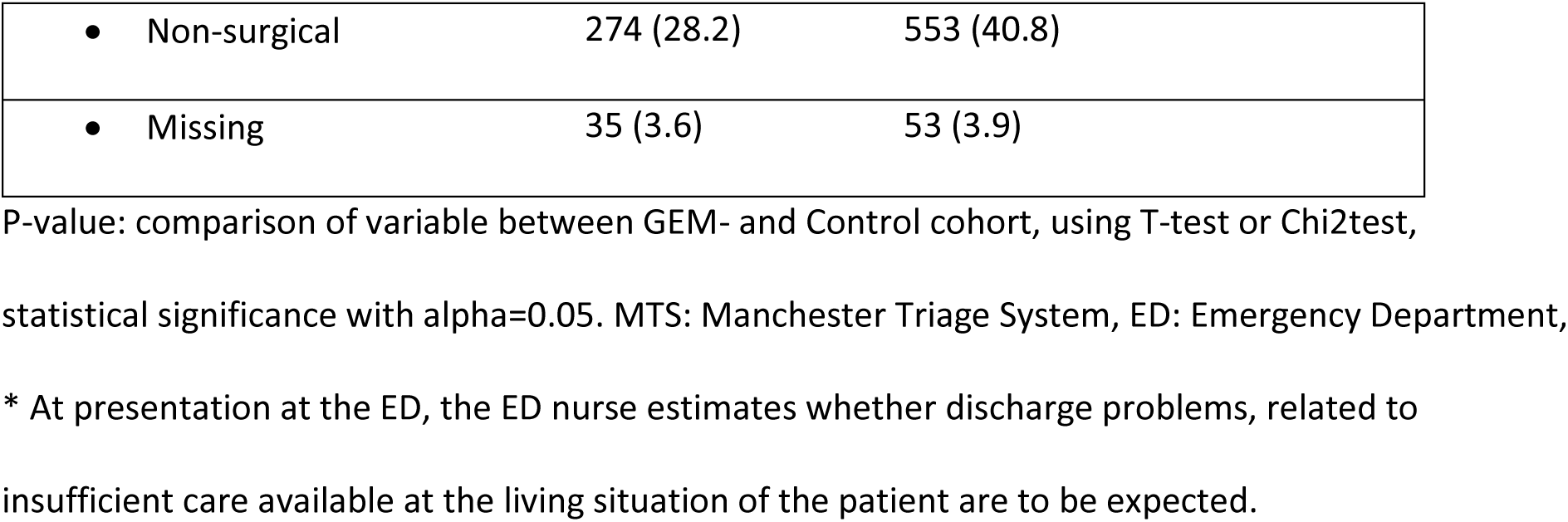
baseline characteristics of all patients.

| Patient characteristics | GEM-cohort<br>n=972 | Control-cohort<br>n=1355 | P-value |
| --- | --- | --- | --- |
| Age in years, mean (SD) | 83.3 (7.0) | 82.7 (7.0) | 0.031 |
| Gender (female), n (%) | 594 (61.1) | 818 (60.4) | 0.718 |
| Triage priority level (MTS), n (%) |  |  | <0.001 |
| • Immediate/<br>Very Urgent | 91 (9.4) | 186 (13.7) |  |
| • Urgent | 351 (36.1) | 537 (39.6) |  |
| • Standard/<br>Non-Urgent | 530 (54.5) | 632 (46.6) |  |
| Presented at the ED between<br>9AM and 5PM on weekdays, n<br>(%) | 909 (93.5) | 564 (41.6) | <0.001 |
| Cognitive impairment, n (%) | 309 (31.8) | 404 (29.8) | 0.308 |
| (High risk of) Delirium, n (%) | 242 (24.9) | 359 (26.5) | 0.385 |
| Presented at ED with<br>Fall/collapse, n (%) | 611 (62.9) | 715 (52.8) | <0.001 |
| Expected discharge problems*,<br>n (%) | 537 (55.2) | 787 (58.1) | 0.173 |
| ED nurse indicates Geriatric Co-management at the ED, n (%) | 638 (65.6) | 896 (66.1) | 0.807 |
| Treating physician surgical/non-surgical n (%) |  |  | <0.001 |
| • Surgical | 663 (68.2) | 749 (55.3) |  |

|  |  |  |
| --- | --- | --- |
| • Non-surgical | 274 (28.2) | 553 (40.8) |
| • Missing | 35 (3.6) | 53 (3.9) |

P-value: comparison of variable between GEM- and Control cohort, using T-test or Chi2test, statistical significance with $\alpha=0.05$ . MTS: Manchester Triage System, ED: Emergency Department, \* At presentation at the ED, the ED nurse estimates whether discharge problems, related to insufficient care available at the living situation of the patient are to be expected.

### Outcomes

The primary outcome of the analyses was the odds of being acutely admitted to the hospital. Secondary outcomes were a composite of 30-day ED readmission and mortality to adjust for competing risks, 90-day mortality and hospital length of stay.

### Analysis

This observational pragmatic study utilized all available data. Baseline categorical data are presented as frequencies and percentages, continuous variables with mean and standard deviation. Baseline differences were calculated using the chi-square test for categorical data and T-test for continuous variables.

To weigh the samples in each group so that the distributions of pre-intervention characteristics were similar between the two groups (34), inverse probability weighting (IPW) was used using the twang package in R (35). A generalized boosted model was used to estimate propensity scores and weights, including baseline covariates as age, gender, triage priority level, primary ED-physician, presence of cognitive problems, delirium, fall/collapse-related presentation, anticipated discharge problems and presence of an indication for geriatric co-management by the ED nurse. To analyze the primary and secondary outcomes, logistic regression models were constructed with co-management by the GEM-team as an independent variable and the propensity score as a covariate. Linear regression was used for hospital length of stay.

### Patient and public involvement

The implementation process of the GEM-team contained analyses of patient journeys and conducting 18 patient and next-of-kin interviews pre- and post-implementation. These were not analyzed in this article. The local patient advisory board was consulted. The outcomes were discussed by the GEM-team and the hospital management. Action items were implemented throughout implementation.

## RESULTS

Approximately 7533 patients aged 70 or older presented at the Emergency Department during the study time. A total of 3932 (52.2%) patients were screened with the GEM-score and 2327 scored positively (Figure 2).

### Descriptive characteristics

Table 1 describes the baseline characteristics of the GEM-cohort and control-cohort. The GEM-cohort patients are older and more likely to present with a fall/collapse at the ED. The Triage priority level according to the Manchester Triage System was overall lower for the GEM-cohort. The GEM-cohort was largely seen during weekdays between 9AM and 5PM and the primary ED-physician was more often a surgical doctor. The average Clinical Frailty Score for the GEM-cohort was five.

### Primary outcome

After adjusting for the propensity score, geriatric co-management at the ED for older patients was associated with decreased odds (odds ratio [OR] 0.61 (95% confidence interval [CI]: 0.50-0.75, P-value <0.001) of hospital admission compared to the control-cohort.

### Secondary outcomes

The composite outcome 30-day ED readmissions and the competing risk of 30-day mortality was not associated with decreased adjusted odds 0.95 (95% CI: 0.75-1.22) compared to the control-cohort. The remaining secondary outcomes are listed in Table 2.

**Table 2.**
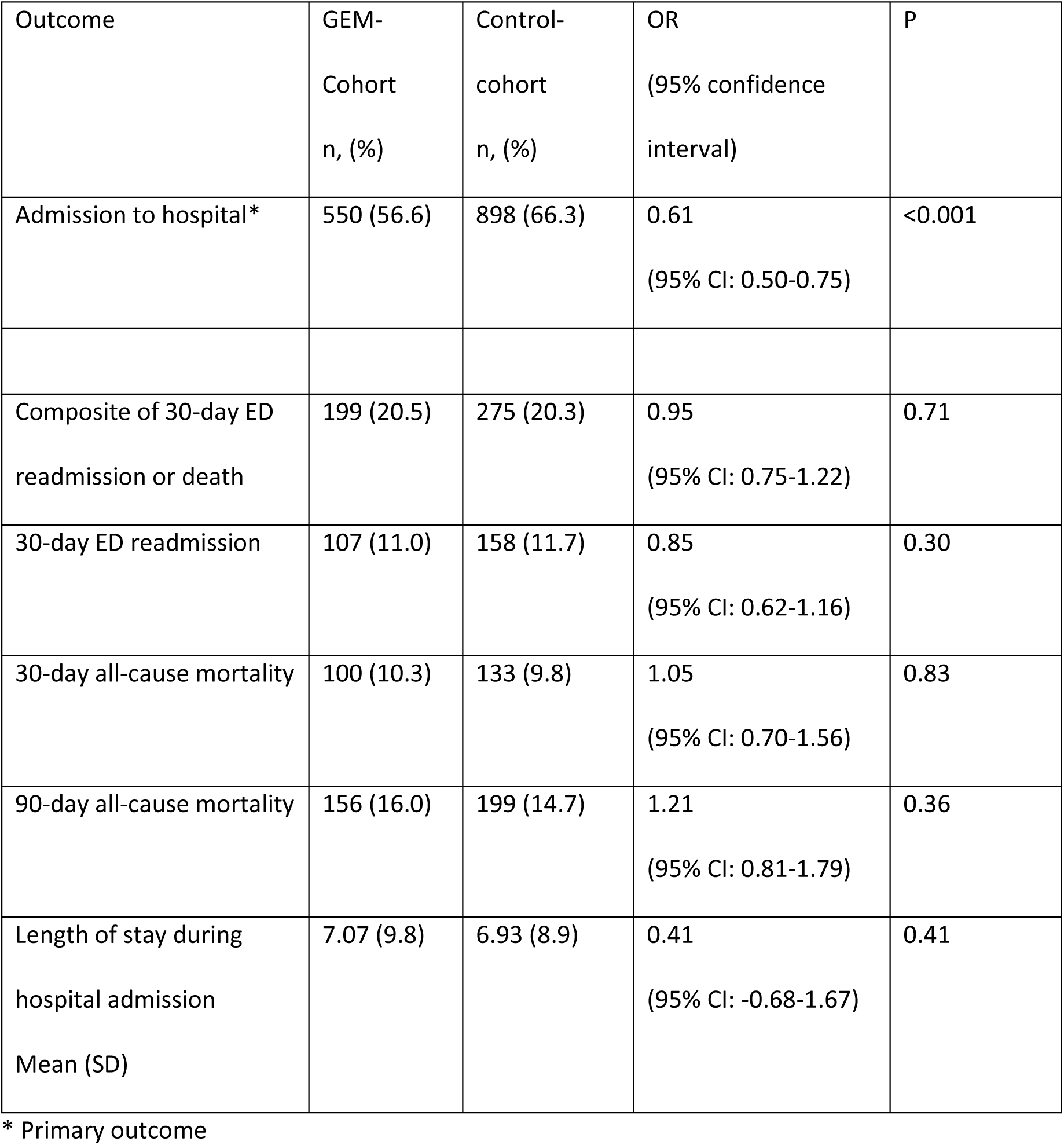
Outcomes of patients co-managed at the Emergency Department by the GEM-team compared to the control-cohort.

| Outcome | GEM-Cohort<br>n, (%) | Control-cohort<br>n, (%) | OR<br>(95% confidence<br>interval) | P |
| --- | --- | --- | --- | --- |
| Admission to hospital* | 550 (56.6) | 898 (66.3) | 0.61<br>(95% CI: 0.50-0.75) | <0.001 |
| Composite of 30-day ED<br>readmission or death | 199 (20.5) | 275 (20.3) | 0.95<br>(95% CI: 0.75-1.22) | 0.71 |
| 30-day ED readmission | 107 (11.0) | 158 (11.7) | 0.85<br>(95% CI: 0.62-1.16) | 0.30 |
| 30-day all-cause mortality | 100 (10.3) | 133 (9.8) | 1.05<br>(95% CI: 0.70-1.56) | 0.83 |
| 90-day all-cause mortality | 156 (16.0) | 199 (14.7) | 1.21<br>(95% CI: 0.81-1.79) | 0.36 |
| Length of stay during<br>hospital admission<br>Mean (SD) | 7.07 (9.8) | 6.93 (8.9) | 0.41<br>(95% CI: -0.68-1.67) | 0.41 |

\* Primary outcome

### Outcomes specifically for the GEM-cohort

The discharge destinations of the GEM-team cohort are summarized in Figure 3, to illustrate that actual transfers to an intermediate or permanent care facility or home did take place, resulting in the lower odds for hospital admission. For the control cohort the discharge destinations are not known other than admitted to the hospital or not.

**Figure 3.**
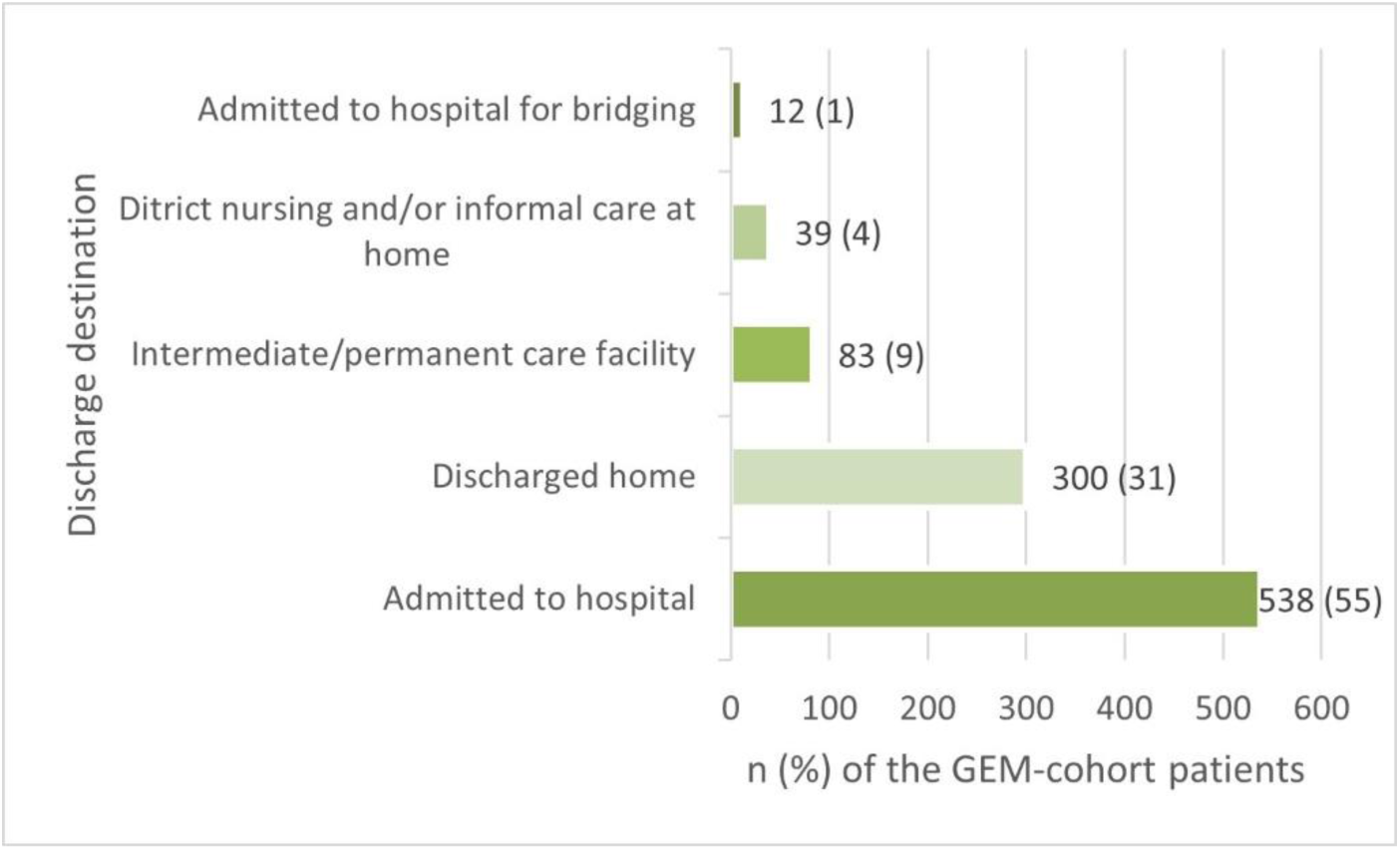
Discharge destinations GEM-cohort: n (%).

## DISCUSSION

This study found that geriatric co-management at the ED is associated with decreased hospital admissions, without having a negative effect on the 30-day ED readmission rates, mortality within 30 or 90 days. These preliminary findings show that the concept of geriatric co-management can effectively be transferred to the ED and result in beneficial outcomes as lower admission to hospital rates without putting the patient at risk for relevant negative outcomes. Examples of intervention components are recognition of non-specific complaints and atypical presentation of illness, attention for relevant co-morbidities, robust medication reviews(32), consistent attention for advance care directives and patient preferences and ongoing working relations with the regional care partners (11, 36, 37).

These findings are like an Australian study where a multidisciplinary team performed targeted geriatric assessment and focused on streamlining of care at the ED (15). The Australian intervention design has large similarities to our geriatric co-management model, however primary assessment was done by an ED-physician alone and selection of the patients was left to the discretion of the multidisciplinary team. No pharmacy team was involved. Geriatric co-management is expected to bring shared responsibility for treatment plans, added resources and assistance, geriatric expertise and working relations with regional care partners (26). By adding this right from primary assessment for a selected group of patients, it is thought that consistent care for a large group of older patients can be delivered. Leaving the responsibility of an important part of the trajectory at the ED with the ED-physician might create dilemmas in prioritizing important but not life-threatening care. In the United States of America, where dedicated Geriatric EDs is increasing based on Geriatric ED guidelines (38), similar outcomes to our study are found. However, in our intervention it was chosen to create a team that relies on working relations with regional care partners. The Dutch healthcare system has a stronger primary care coordination system compared to the USA (39) and Dutch governmental health care bodies stimulate regional collaboration as a strategy to keep the healthcare system accessible.

In contrast to the outcomes of a systematic review suggesting that multi-strategy interventions and a large follow-up may be associated with a reduction of hospital admission and 30-day ED readmissions(19), geriatric co-management did not result in lower risk for 30-day ED readmissions. This might be related to the short time of the intervention only during the ED presentation and no long term follow up.

### Strengths and limitations

The design of this observational study enabled us to analyze outcomes of all older patients presented at the ED representing a real-world population. This contributes to the feasibility of geriatric co-management at the ED. As the start of the intervention and data inclusion coincided with the first SARS-CoV-2 epidemic period possibly affecting outcomes to other differences, a control cohort out of the same time window was created. Although the ED-nurses were instructed to apply the score to all aged 70 or older, only 52% of the patients were screened with the GEM score. Especially in the out of office hours it proved difficult to incorporate the GEM-score in the regular work processes.

The GEM-team was available on weekdays between 9 AM and 5 PM. 94% of the GEM-cohort was seen between these hours versus 42% of the control-cohort, this is a large baseline difference. It was not chosen to just compare those presented under office hours, as those who were not seen by the GEM-team during office hours, were judged by routine practice at the ED, considered less frail and/or having a low chance of unnecessary hospital admission, resulting in a bias in favor of this potential control-group. The 58% of the control-cohort that was presented outside the working hours of the GEM-team could have had a higher risk for hospital admission related to being sicker and in need of ED presentation which could not wait until the next working day. To control this baseline imbalance, a propensity score was created, including triage priority level, presence of cognitive problems, delirium, falls and anticipated discharge problems. The triage priority level represents an assessment of the severity of disease and an interpretation of the vital signs. Also, the control-cohort patients had access to the regional covenant protocol which made it possible to transfer ED-patients to an intermediate care facility at all hours. However, some confounding related to the hours of presentation cannot be fully excluded. Therefore, this study, by choosing an observational study design, limits itself to assess an association and not a causal relation. The external validity of this study, bridging the evidence to implementation gap is high. However, this study is limited in assessing the GEM-team’s internal validity(40). The anonymized data available to this study was limited and much depending on regular hospital systems and possibilities within privacy regulations. Due to a change in hospital systems, it wasn’t possible to analyze length of stay at the ED. The strength of the collaboration of the GEM-team and the regional network is of influence on the generalizability of these study results.

### Recommendations

Geriatric co-management was already seen as a promising approach to hospital care(41). These preliminary study results could imply that geriatric co-management at the ED might be of interest too. It contributes to an integrated healthcare system approach, which we are going to need to keep our healthcare system accessible during the demographic changes heading our way in the coming years(42). In the Netherlands it is of high priority to only use hospital beds for those who need and desire treatment. As the GEM-team helps to select the right patients for transmission to home or an intermediate or long-term care facilities, it is possible to deliver more care without the need for more expensive clinical beds. Presentation to the ED by patients living with frailty cannot always be avoided as quick and urgent clinical assessments might be indicated, but admission to hospital without a need or desire for treatment can be prevented.

Future studies would be to evaluate the GEM-team and its association on quality of life, patient experiences and patient related outcomes. Multi-center studies with clustered RCT cross-over designs could be considered. The GEM-score needs to be evaluated and perspectives on the implementation process might be of benefit to generalize these results. It would be of interest to evaluate the effectiveness of geriatric co-management at the ED in other healthcare systems.

## Conclusion

Geriatric co-management at the ED is associated with decreased hospital admissions while 30-day ED readmissions or mortality was not impacted. These preliminary results contribute to the evidence that geriatric co-management may be an effective intervention for older patients with frailty at the ED.

## DECLARATIONS

### Availability of data and materials

The datasets used and/or analyzed during the current study are available from the corresponding author on reasonable request.

### Authors Contributions

Conceptualization: VH, AdJ; methodology: VH, MR, JMV; investigation: VH, MR, JMV writing—original draft preparation, VH; writing—review and editing: VH, JMV, MR, AdJ, BB; supervision, JMV, AdJ, BB. All authors have read and agreed to the published version of the manuscript.

## Data Availability

All data produced in the present study are available upon reasonable request to the authors

## Additional file 1

STROBE statement – checklist of items that should be included in reports of cohort studies

The TIDieR checklist, Information to include when describing an intervention and the location of the information.

