## Additional file 1 for "Association between older patients receiving geriatric co-management at the emergency department and acute hospital admissions compared to usual care: an observational, controlled study"

STROBE Statement—Checklist of items that should be included in reports of ***cohort studies***

|  | Item No | Recommendation | Page No |
| --- | --- | --- | --- |
| **Title and abstract** | 1 | (*a*) Indicate the study’s design with a commonly used term in the title or the abstract | 1-3 |
|  |  | (*b*) Provide in the abstract an informative and balanced summary of what was done and what was found |  |
| Introduction | | | |
| Background/rationale | 2 | Explain the scientific background and rationale for the investigation being reported | 5-6 |
| Objectives | 3 | State specific objectives, including any prespecified hypotheses | 6 |
| Methods | | | |
| Study design | 4 | Present key elements of study design early in the paper | 7 |
| Setting | 5 | Describe the setting, locations, and relevant dates, including periods of recruitment, exposure, follow-up, and data collection | 7-10 |
| Participants | 6 | (*a*) Give the eligibility criteria, and the sources and methods of selection of participants. Describe methods of follow-up | 7-9 |
|  |  | (*b*) For matched studies, give matching criteria and number of exposed and unexposed |  |
| Variables | 7 | Clearly define all outcomes, exposures, predictors, potential confounders, and effect modifiers. Give diagnostic criteria, if applicable | 10 |
| Data sources/ measurement | 8* | For each variable of interest, give sources of data and details of methods of assessment (measurement). Describe comparability of assessment methods if there is more than one group | 10 |
| Bias | 9 | Describe any efforts to address potential sources of bias | 11 |
| Study size | 10 | Explain how the study size was arrived at | 11 |
| Quantitative variables | 11 | Explain how quantitative variables were handled in the analyses. If applicable, describe which groupings were chosen and why | - |
| Statistical methods | 12 | (*a*) Describe all statistical methods, including those used to control for confounding | 10 |
|  |  | (*b*) Describe any methods used to examine subgroups and interactions |  |
|  |  | (*c*) Explain how missing data were addressed |  |
|  |  | (*d*) If applicable, explain how loss to follow-up was addressed |  |
|  |  | (*e*) Describe any sensitivity analyses |  |
| Results | | |  |
| Participants | 13* | (a) Report numbers of individuals at each stage of study—eg numbers potentially eligible, examined for eligibility, confirmed eligible, included in the study, completing follow-up, and analysed | 12 |
|  |  | (b) Give reasons for non-participation at each stage |  |
|  |  | (c) Consider use of a flow diagram |  |
| Descriptive data | 14* | (a) Give characteristics of study participants (eg demographic, clinical, social) and information on exposures and potential confounders | 12-13 |
|  |  | (b) Indicate number of participants with missing data for each variable of interest |  |
|  |  | (c) Summarise follow-up time (eg, average and total amount) |  |
| Outcome data | 15* | Report numbers of outcome events or summary measures over time | 15 |

| Main results | 16 | (*a*) Give unadjusted estimates and, if applicable, confounder-adjusted estimates and their precision (eg, 95% confidence interval). Make clear which confounders were adjusted for and why they were included | 14-15 |
| --- | --- | --- | --- |
|  |  | (*b*) Report category boundaries when continuous variables were categorized |  |
|  |  | (*c*) If relevant, consider translating estimates of relative risk into absolute risk for a meaningful time period |  |
| Other analyses | 17 | Report other analyses done—eg analyses of subgroups and interactions, and sensitivity analyses | - |
| Discussion | | | |
| Key results | 18 | Summarise key results with reference to study objectives | 17 |
| Limitations | 19 | Discuss limitations of the study, taking into account sources of potential bias or imprecision. Discuss both direction and magnitude of any potential bias | 18-19 |
| Interpretation | 20 | Give a cautious overall interpretation of results considering objectives, limitations, multiplicity of analyses, results from similar studies, and other relevant evidence | 19-20 |
| Generalisability | 21 | Discuss the generalisability (external validity) of the study results | 19-20 |
| Other information | | | |
| Funding | 22 | Give the source of funding and the role of the funders for the present study and, if applicable, for the original study on which the present article is based | 22 |

*Give information separately for exposed and unexposed groups.

**Note:** An Explanation and Elaboration article discusses each checklist item and gives methodological background and published examples of transparent reporting. The STROBE checklist is best used in conjunction with this article (freely available on the Web sites of PLoS Medicine at http://www.plosmedicine.org/, Annals of Internal Medicine at http://www.annals.org/, and Epidemiology at http://www.epidem.com/). Information on the STROBE Initiative is available at http://www.strobe-statement.org.

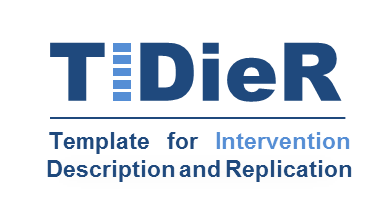

**The TIDieR (Template for Intervention Description and Replication) Checklist*:**

Information to include when describing an intervention and the location of the information

| **Item number** | **Item** | **Where located **** | |
| --- | --- | --- | --- |
|  |  | Primary paper  (page or appendix  number) | Other ^†^ (details) |
|  | **BRIEF NAME** |  |  |
| **1.** | Provide the name or a phrase that describes the intervention. | _______1____ | ______________ |
|  | **WHY** |  |  |
| **2.** | Describe any rationale, theory, or goal of the elements essential to the intervention. | _____6-7___ | _____________ |
|  | **WHAT** |  |  |
| **3.** | Materials: Describe any physical or informational materials used in the intervention, including those provided to participants or used in intervention delivery or in training of intervention providers. Provide information on where the materials can be accessed (e.g. online appendix, URL). | ____8-14_____ | _____________ |
| **4.** | Procedures: Describe each of the procedures, activities, and/or processes used in the intervention, including any enabling or support activities. | ____8-11_____ | _____________ |
|  | **WHO PROVIDED** |  |  |
| **5.** | For each category of intervention provider (e.g. psychologist, nursing assistant), describe their expertise, background and any specific training given. | ____9________ | _____________ |
|  | **HOW** |  |  |
| **6.** | Describe the modes of delivery (e.g. face-to-face or by some other mechanism, such as internet or telephone) of the intervention and whether it was provided individually or in a group. | _____8-11______ | _____________ |
|  | **WHERE** |  |  |
| **7.** | Describe the type(s) of location(s) where the intervention occurred, including any necessary infrastructure or relevant features. | ______8_______ | _____________ |
|  | **WHEN and HOW MUCH** |  |  |
| **8.** | Describe the number of times the intervention was delivered and over what period of time including the number of sessions, their schedule, and their duration, intensity or dose. | ______15_____ | _____________ |
|  | **TAILORING** |  |  |
| **9.** | If the intervention was planned to be personalised, titrated or adapted, then describe what, why, when, and how. | _______N/A____ | _____________ |
|  | **MODIFICATIONS** |  |  |
| **10.^ǂ^** | If the intervention was modified during the course of the study, describe the changes (what, why, when, and how). | _______N/A___ | _____________ |
|  | **HOW WELL** |  |  |
| **11.** | Planned: If intervention adherence or fidelity was assessed, describe how and by whom, and if any strategies were used to maintain or improve fidelity, describe them. | ______N/A_____ | _____________ |
| **12.^ǂ^** | Actual: If intervention adherence or fidelity was assessed, describe the extent to which the intervention was delivered as planned. | _____N/A______ | _____________ |

** **Authors** - use N/A if an item is not applicable for the intervention being described. **Reviewers** – use ‘?’ if information about the element is not reported/not sufficiently reported.

† If the information is not provided in the primary paper, give details of where this information is available. This may include locations such as a published protocol or other published papers (provide citation details) or a website (provide the URL).

ǂ If completing the TIDieR checklist for a protocol, these items are not relevant to the protocol and cannot be described until the study is complete.

* We strongly recommend using this checklist in conjunction with the TIDieR guide (see *BMJ* 2014;348:g1687) which contains an explanation and elaboration for each item.

* The focus of TIDieR is on reporting details of the intervention elements (and where relevant, comparison elements) of a study. Other elements and methodological features of studies are covered by other reporting statements and checklists and have not been duplicated as part of the TIDieR checklist. When a **randomised trial** is being reported, the TIDieR checklist should be used in conjunction with the CONSORT statement (see [www.consort-statement.org](http://www.consort-statement.org)) as an extension of **Item 5 of the CONSORT 2010 Statement.** When a **clinical trial** **protocol** is being reported, the TIDieR checklist should be used in conjunction with the SPIRIT statement as an extension of **Item 11 of the SPIRIT 2013 Statement** (see [www.spirit-statement.org](http://www.spirit-statement.org)). For alternate study designs, TIDieR can be used in conjunction with the appropriate checklist for that study design (see [www.equator-network.org](http://www.equator-network.org)).
